# Genome-wide association study of liking of physical activity in the UK Biobank

**DOI:** 10.1101/2021.10.13.21264969

**Authors:** Yann C. Klimentidis, Michelle Newell, Matthijs D. van der Zee, Victoria L. Bland, Sebastian May-Wilson, Cristina Menni, Massimo Mangino, Amit Arora, David A. Raichlen, Gene E. Alexander, James F. Wilson, Dorret I. Boomsma, Jouke-Jan Hottenga, Eco J.C. de Geus, Nicola Pirastu

## Abstract

A lack of physical activity (PA) is one of the most pressing health issues facing society today. Our individual propensity for PA is partly influenced by genetic factors. Stated liking of various PA behaviors may capture additional dimensions of PA behavior that are not captured by other measures, and contribute to our understanding of the genetics of PA behavior. Here, in over 157,000 individuals from the UK Biobank, we sought to complement and extend previous findings on the genetics of PA behavior by performing genome-wide association studies of self-reported liking of several PA-related behaviors plus an additional derived trait of overall PA-liking. We identified a total of 19 unique genome-wide significant loci across all traits, only four of which overlap with loci previously identified for PA behavior. The PA-liking traits were genetically correlated with self-reported (r_g_: 0.38 – 0.80) and accelerometry-derived (r_g_: 0.26 – 0.49) PA measures, and with a wide range of health-related traits and dietary behaviors. Replication in the Netherlands Twin Register (NTR; n>7,300) and the TwinsUK (n>1,300) study revealed directionally consistent associations. Polygenic risk scores (PRS) were then trained in UKB for each PA-liking trait and for self-reported PA behavior. The PA-liking PRS significantly predicted the same liking trait in NTR. The PRS for liking of *going to the gym* predicted PA behavior in NTR (r^2^ = 0.40%) nearly as well as the one constructed based on self-reported PA behavior (r^2^ = 0.42%). Combining the two PRS into a single model increased the r^2^ to 0.59%, suggesting that although these PRS correlate with each other, they are also capturing distinct dimensions of PA behavior. In conclusion, we have identified the first loci associated with PA-liking, and extended and refined our understanding of the genetic basis of PA behavior.

## INTRODUCTION

Levels of physical activity (PA) have decreased dramatically in most parts of the world over the past several hundred years, likely contributing to a major and growing chronic disease burden ^1–3^. Physical inactivity has been compared to smoking in terms of its impact on disease burden, which ranges widely from cardiometabolic disease to mental health ^1^. As genetic factors partly explain individual differences in PA behavior ^4–7^, identifying specific genetic risk factors can advance our understanding of 1) important inter-individual variation, 2) relevant biological pathways, and 3) the presence, direction, and strength of causal relationships between PA behaviors and health outcomes.

Several loci have already been identified as being associated with self-reported and accelerometry-measured levels of PA ^8,9^. These measures may each be limited in multiple ways. For example, self-reported measures may be highly influenced by social- and health-related pressures and may not be stable over time, while accelerometer measures may only be sensitive to certain types of PA, with wear-time often limited to a single week of a person’s lifetime and likely influencing behavior ^10^. Measures of an individual’s liking of PA may more accurately capture overall life-long propensity to engage in PA, and at a minimum, serve as complementary, broader, and refined measures of PA behavior.

To discover genetic loci associated with PA-related liking, we performed GWAS of five individual liking traits and one composite trait in over 157,000 UK Biobank participants, with replication of top loci in the Netherlands Twin Register (NTR) and TwinsUK studies. We then examined how these traits genetically relate to self-reported and accelerometry-measured PA ^8^, and to a wide range of other traits and health outcomes. Finally, we examined how polygenic scores (PGS) of PA-liking derived from the UK Biobank predicted both liking and self-reported PA in the NTR study.

## METHODS

### UK Biobank

The UK Biobank is a prospective cohort study of 500,000 adults (ages 37-73 at the baseline examination in 2006-2010) from the UK ^11^. All participants provided written informed consent, and ethical approval for this study was granted. Ethical approval for the UK Biobank study was obtained by the National Information Governance Board for Health and Social Care, and the National Health Service North West Multicenter Research Ethics Committee.

### Physical activity liking

In 2019, a link to a questionnaire was sent by email to UK Biobank participants to assess liking of specific foods as well as physical activities ^12^. This online questionnaire consisted of 150 items, five of which were related to PA (*going to the gym, working up a sweat, exercising with others, exercising alone*, and *bicycling*), and assesses liking through a 9-point hedonic scale ranging from 1 for extremely dislike up to 9 for extremely like, in increments of one. The same items were asked in the two replication studies (see below).

### Genetic markers

Genotypes in the UK Biobank were measured with the Affymetrix UK Biobank Axiom Array (Santa Clara, CA, USA) in 90% of participants. The remainder (10%) were genotyped with the Affymetrix UK BiLEVE Axiom Array. Further details about imputation, principal components analysis, and QC procedures can be found elsewhere ^13^.

### Replication of sentinel SNPs in Netherlands Twin Register (NTR) and TwinsUK

The NTR is a longitudinal register of twins and their relatives ^14^. Between December 2014 and May 2017, participants responded to the same liking questionnaire as the one administered in the UK Biobank, including the same 5 PA-related questions ^15^. Details regarding genotyping are provided in Supplementary Methods. Top genome-wide significant SNPs or their proxies from the UK Biobank GWASs were interrogated for each respective trait in NTR GWAS results. Since we tested 25 SNPs/loci, a replication was deemed successful if the p-value was < 0.002. Polygenic scores (PGS) were calculated in NTR with the LDPRED package ^16^, based on UKB GWAS results for PA-liking and for SSOE (strenuous sports or other exercise) and other PA behavior measures ^8^, and were tested for correlation with PA-liking phenotypes and self-reported PA behavior in NTR (see Supplementary Methods for more details).

TwinsUK is a large twin registry for the study of health that began in 1992 ^17^. The same liking questionnaire used in UK Biobank was previously used in the TwinsUK cohort ^18^. TwinsUK genotype has been previously described in detail ^19^. Briefly, TwinsUK samples were genotyped with a combination of two Illumina arrays (HumanHap300, HumanHap610Q). After the Genotype QC stage, the samples from the two arrays were combined and the imputations were performed using the Michigan Imputation Server ^20^ using the 1000 Genomes Phase3 v5 reference panel.

### Statistical analyses

To assess associations of PA liking with sex, age, body mass index (BMI), income, University/College degree (yes/no), and Townsend Deprivation Index, we performed linear regression after ensuring normality and homoscedasticity of residuals. In GWAS, we included only individuals of European descent. We considered participants as being of European descent if they were either among the genetically British as defined by UK Biobank or self-identified as “Irish”, “White”, or “Any other White background”. We performed GWAS with fastGWA ^21^, which implements a mixed-effect linear regression that controls for population stratification and relatedness. We included age at time of questionnaire, sex, genotyping chip, batch, and the first 10 genetic principal components, as covariates. We used LD-score regression to assess test score inflation, SNP-based heritability, and to assess genetic correlations among PA-liking traits and previously reported PA traits ^22^. To obtain a measure of overall PA liking, we also derived a GWAS of the first principal component (PC) derived through the genetically-independent phenotype (GIP) method ^23,24^ and starting from the genetic correlation matrix to derive the loadings of each trait on each GIP. Independent significant loci were identified as those with p < 5 × 10^−8^ with r^2^ < 0.1, and > 250 kb distance. The online LDHUB platform ^25^ was used to examine genetic correlations of the five individual liking traits and the overall PA-liking (GIP1) trait with a wide range of traits and diseases (∼ 800 phenotypes). We used stratified LD score regression to identify cell type-specific enrichment of heritability ^26^ from the overall PA-liking (GIP1) GWAS.

## RESULTS

### PA-liking phenotypes

Descriptive statistics of UKB, NTR, and TwinsUK samples and mean PA-liking levels are shown in **Supplementary Table 1**. Mean score of liking was highest for *exercising with others* in the two UK samples, and highest for *bicycling* in NTR. It was lowest for *going to the gym* in all three samples. In the UK Biobank, PA liking was negatively correlated with age, and was higher in males (except for *exercising with others*). We observed generally positive correlations of PA-liking traits with education and income, and negative correlations with Townsend Deprivation Index (e.g. *exercising with others*), such that less deprivation was associated with more PA-liking **(Supplementary Table 2)**.

### Genome-wide association study

Data from up to 158,189 UK Biobank participants (mean age=66.8; 57% female) were analyzed for GWAS. SNP-based trait heritabilities varied from 0.054 (0.004) for *going to the gym*, up to 0.075 (0.004) for *bicycling*. SNP heritability for overall PA-liking (GIP1) was 0.089 (0.004) **(Supplementary Table 3)**. We did not observe evidence of genomic inflation beyond that explained by polygenic signal according to LD-score regression estimates **(Supplementary Table 2)**. Between 2 and 6 genome-wide significant loci were identified for each individual liking trait, and 8 loci for overall PA-liking (GIP1), for a total of 25 SNP-trait associations in 19 loci **(Figure 1 and Table 1)**. We did not observe a large degree of overlap of top loci across the five different liking traits. Only two loci (*CADM2* and the locus on chromosome 11) were significantly associated with more than one trait. Sentinel SNPs were also associated with other traits such as with bone mineral density, body size and body composition, educational attainment, respiratory traits, psychiatric traits, as well as other PA-related traits such as usual walking pace and time watching television (**Supplementary spreadsheet**). Cell-type enrichment analysis via stratified LD score regression identified the nucleus accumbens, hippocampus, caudate, frontal cortex, and amygdala cell types **(Supplementary Table 4)**.

**Table 1:**
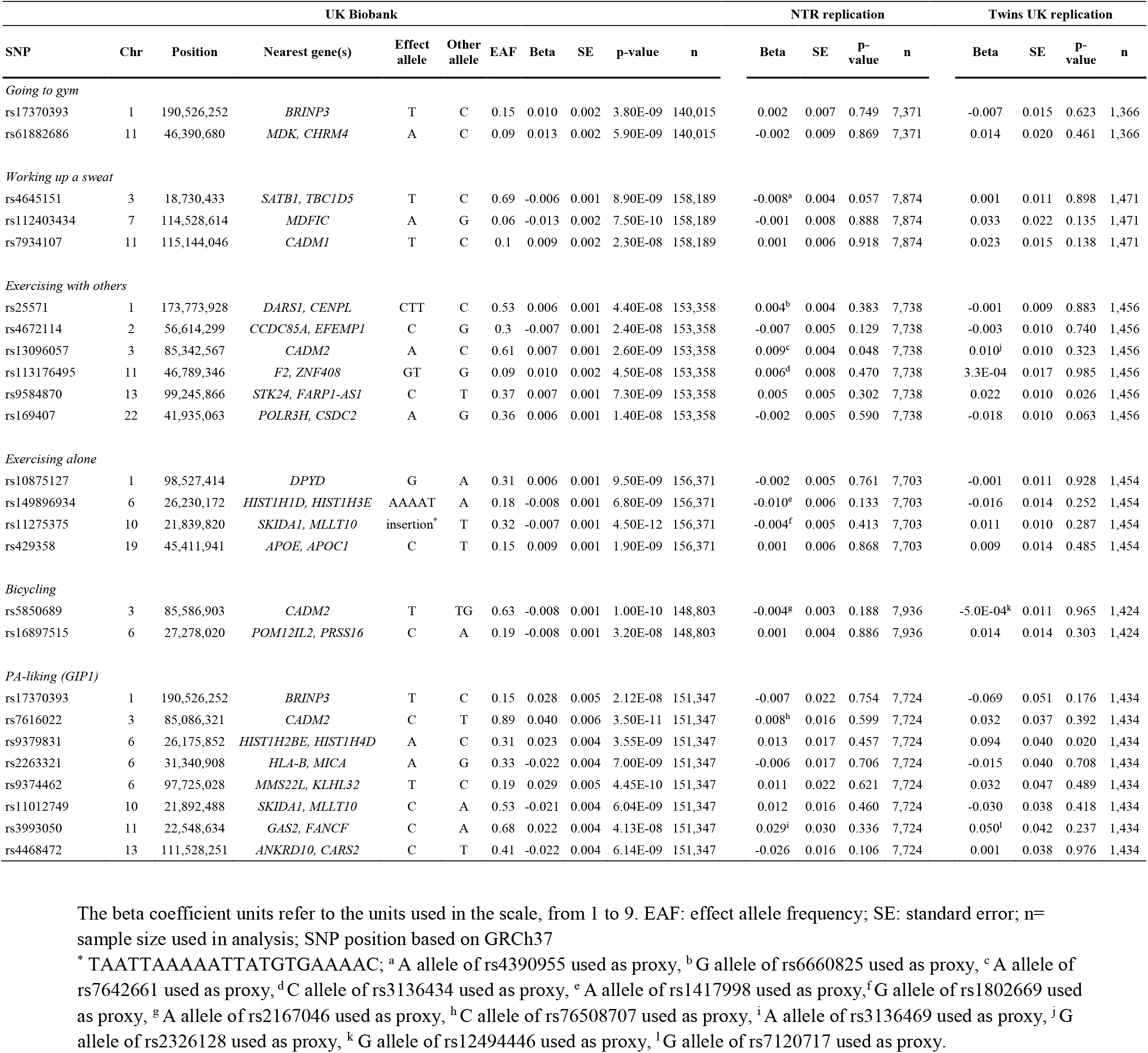
List of genetic variants associated with physical activity (PA) liking and overall PA-liking (GIP1) in UKB, with replication results from NTR and Twins UK.

**Figure 1:**
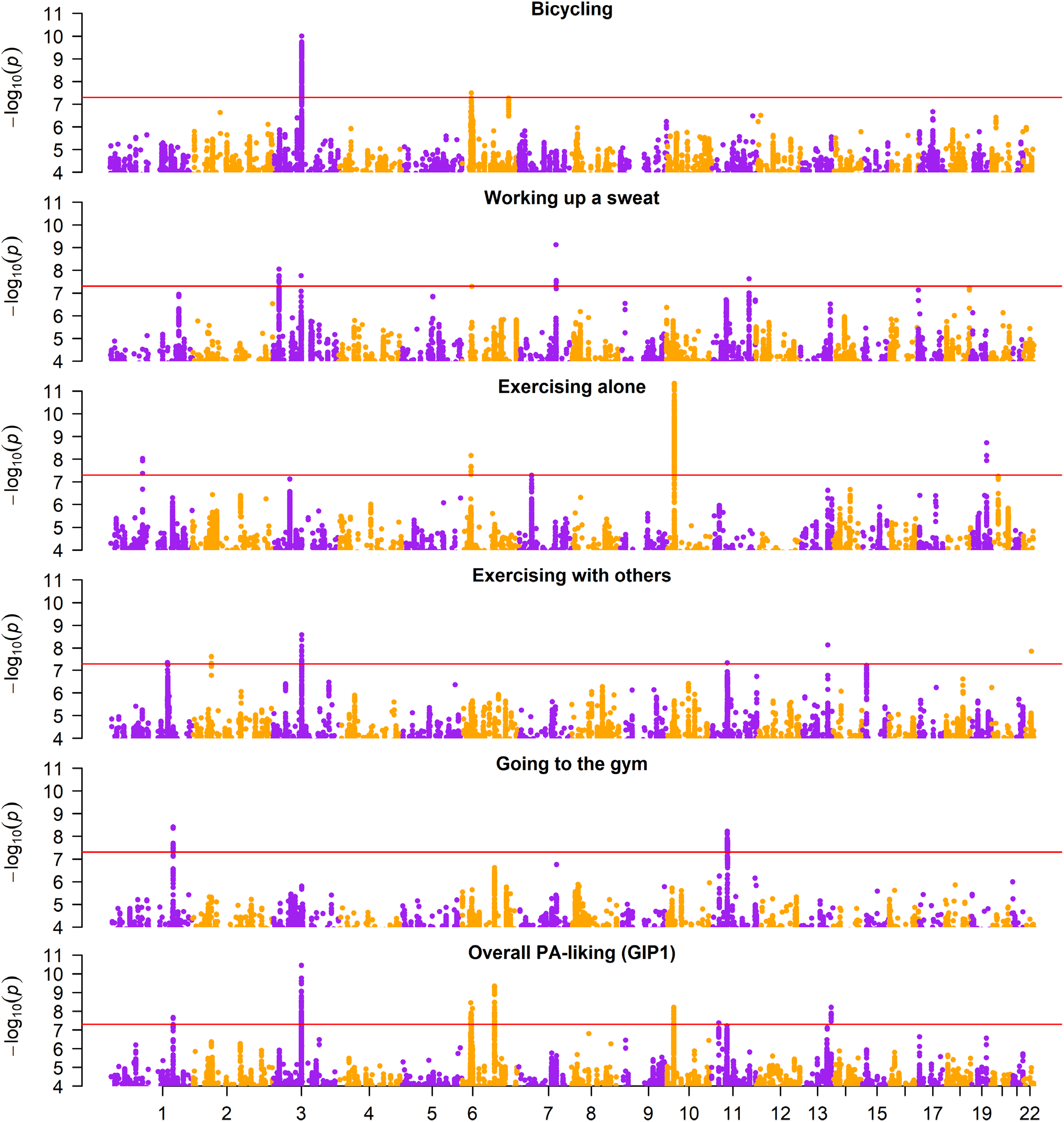
Manhattan plots of five physical activity liking traits and overall PA-liking (GIP1).

In the NTR replication, we found directional consistency for 19 out of 25 SNPs, including 6 out of 8 PA-liking (GIP1) SNPs (**Table 1**). Only one SNP (*CADM2*; for *exercising with others*) was nominally significant (p<0.05) before multiple testing correction. In the TwinsUK replication, we found directional consistency for 14 out of 25 SNPs, including 6 out of 8 overall PA-liking (GIP1) SNPs (**Table 1**).

### Genetic correlations

Genetic correlations among PA liking traits were moderate-to-strong (**Figure 2**). Among the PA liking traits, strong correlations were observed for *working up a sweat* with *going to the gym* (r_g_=0.79), *exercising with others* (r_g_=0.76), and *exercising alone* (r_g_=0.72), and the weakest correlation was a positive one between *exercising alone* and *exercising with others* (r_g_=0.46). Across liking and behavior (self-reported PA and accelerometer-derived PA) traits, correlations were strongest for PA-liking with self-reported vigorous physical activity and strenuous sports and other exercise (r_g_ between 0.59 and 0.78). Genetic correlations of PA-liking with accelerometry traits were generally strongest for *exercising alone* (r_g_≈0.47), and weakest with *going to the gym* (r_g_≈0.28). Genetic correlation assessments with a wide range of traits and diseases reveal correlations with UK Biobank variables related to physical activity including accelerometry, as well as with obesity-related traits, tiredness, and lifestyle traits such as alcohol consumption, TV watching, and taking dietary supplements, among others (**Figure 3**).

**Figure 2:**
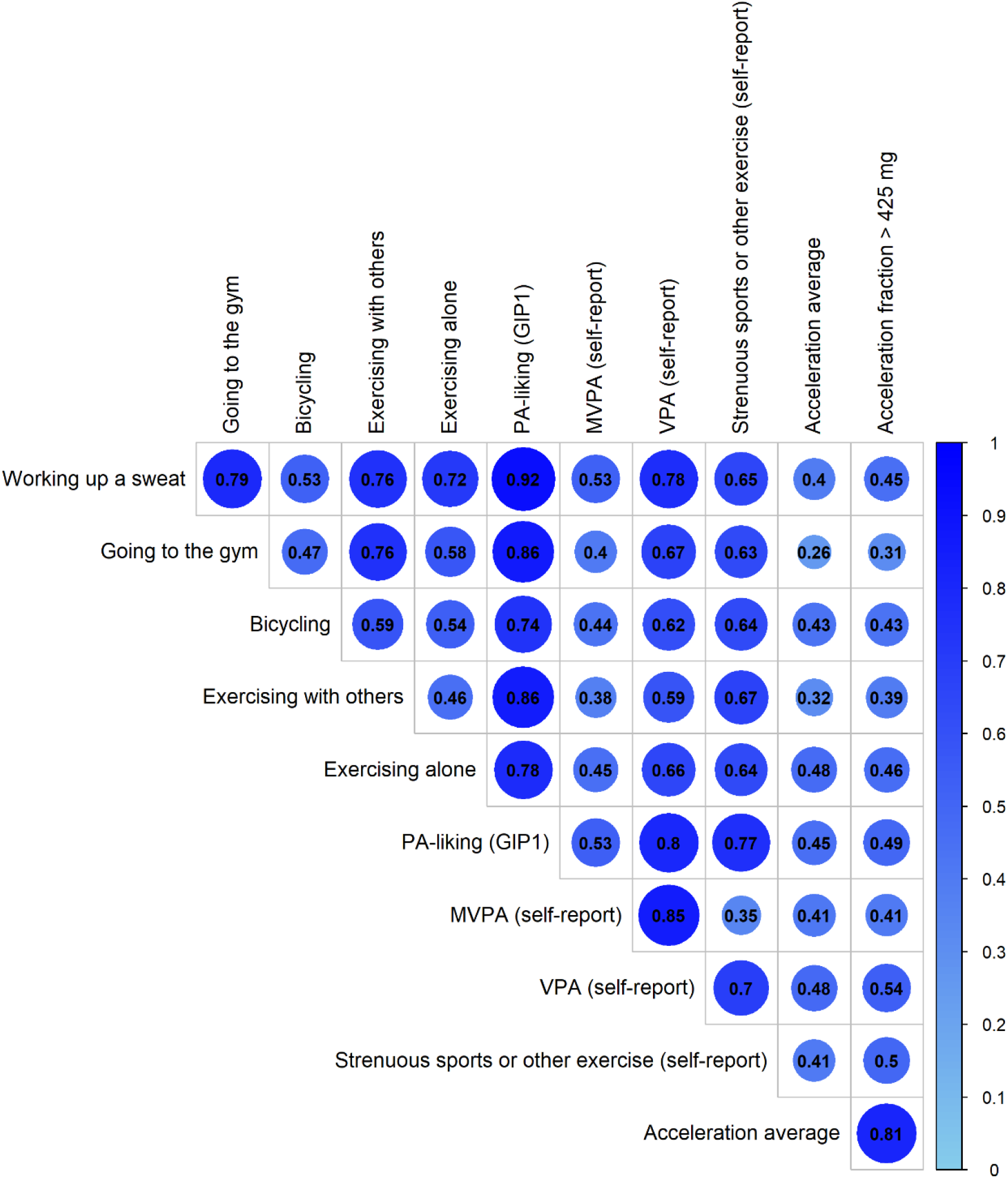
Genetic Correlations across PA self-report, PA from accelerometry, and PA-liking traits.

**Figure 3:**
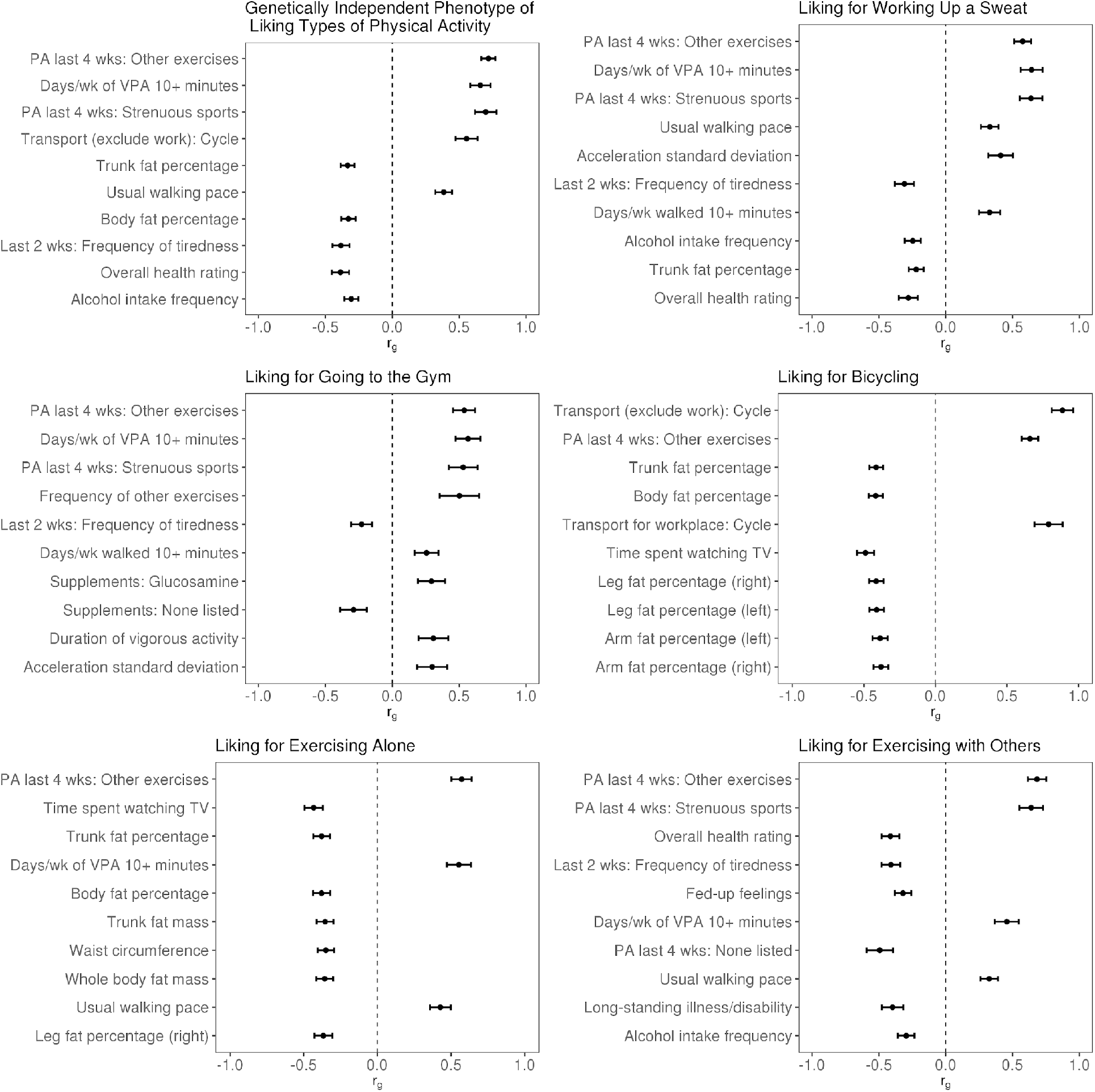
Genetic correlations of PA-liking traits with other traits and diseases. Top ten correlations are shown according to p-value. The order of traits shown in each panel is in ascending order by p-value. Error bars represent 95 % CI.

### PRS analyses in NTR

PRSs of each PA-liking trait were calculated for each NTR participant, using as weights the effect sizes resulting from the GWAS of the UK Biobank data. These PRSs were generally, but not always, most strongly associated with the corresponding PA-liking phenotype (**Supplementary Table 4)**. For example, the *exercising with others* PRS was most strongly associated with *exercising with others* in NTR (r^2^=0.79%, p=2.7 × 10^−13^). The overall PA-liking (GIP1) PRS was most strongly correlated with liking of *exercising alone* (r^2^=0.83%, p=7.0 × 10^−15^; **Supplementary Table 4)**. These PRSs were also significantly associated with self-reported PA behavioral phenotypes in NTR: self-reported *total exercise, team-based exercise*, and *solitary exercise*. Among the PA-liking PRSs, the best predictors of these self-reported PA phenotypes were *going to the gym*, and *exercising with others* in association with the self-reported PA measures of *total exercise* and *solitary exercise* (but not with *team-based exercise*; **Supplementary Figure 1**). When compared with the PRS based on the self-reported and accelerometer PA GWAS in UK Biobank, the liking PRSs had a similar predictive performance. For example, a PRS of self-reported SSOE (strenuous sports or other exercise) predicted self-report *total exercise* in NTR only slightly better than a PRS of liking *going to the gym* (**Supplementary Figure 1**). When the best predicting liking PRS (*going to the gym*) was combined with the best predicting self-reported PA PRS (SSOE), the prediction of self-reported PA in NTR improved from r^2^=0.42% to r^2^=0.59%, corresponding to a 40% improvement in prediction, suggesting that these two measures are capturing distinct and complementary components of PA (**Supplementary Figure 2**).

## DISCUSSION

With the advent of large-scale biobanks with genetic data, we are now able to identify genomic loci associated with complex behavioral and health-related traits such as PA. Here, we broaden and deepen our nascent understanding of the genetics of PA behavior by identifying genetic variants associated with PA-liking. We found some but minimal overlap of loci with those identified for self-reported and accelerometer measures of PA, found genetic correlations with other health-related traits, including behaviors and health outcomes, and find that PRS of PA-liking adds substantially to prediction of PA in an independent sample, beyond a prediction based on a self-reported PA PRS.

SNP-based heritabilities of PA-liking traits (ranging from 5.4 to 7.5 %; PA-liking (GIP1) = 8.9 %) were generally higher than those of self-reported behaviors at the baseline exam (ranging from 4.6 to 5.6 %), but lower than accelerometry measures (14.3 and 11.0 %) ^8^, likely due to lower measurement error of accelerometry. Of all 24 loci identified for liking traits, only *APOE, CADM2, HIST1H1D*, and *SKIDA1* were significantly associated with self-reported or accelerometry-measured PA ^8,9^. However, it is likely that some of the other 20 loci are associated with PA behavior at a less stringent significance threshold. This relatively small degree of overlap likely suggests that these measures of liking are reflecting different and specific facets of PA, and attitudes and perceptions about PA, as opposed to PA behavior assessed by self-report or accelerometry. It is possible that since accelerometers measure overall PA, and not just purposeful exercise, the PA-liking traits are more strongly genetically correlated with self-reported PA than with accelerometer measures of PA. On the other hand, the relatively low genetic correlation of *going to the gym* with accelerometer-measured PA may reflect limitations of accelerometer measurements in the context of certain types of PA such as resistance exercise. Genetic correlation analyses across a wider set of traits and diseases mainly revealed correlations of PA-liking with self-reported PA and body fat measures. However, several other notable findings such as negative genetic correlations with frequency of tiredness, fed-up feelings, and alcohol intake frequency, and positive associations with usual walking pace, supplement intakes, and variance in accelerometer measurements in UKB, were also observed. Some of these correlations may represent causal effects in one or both directions.

Among the individual loci identified, *CADM2* has previously been identified in GWAS of BMI ^27^, risk taking and other behavioral traits ^28–30^, including PA ^8^. However, the pattern of association with *CADM2* variants is particularly interesting since alleles associated with higher BMI are associated with *higher* levels of PA and PA-liking, in the opposite direction of the phenotypic association. As the most consistently identified genetic locus across self-reported PA and PA-liking measures, we will benefit from future work to understand the molecular mechanisms underlying this association. We found a SNP (rs7934107-T) in another cell adhesion molecule gene, *CADM1*, which was associated with *working up a sweat*. A SNP in the same gene was found to be associated with anorexia nervosa (SNP-risk allele: rs6589488-A) ^31^. These alleles at these two SNPs are positively and moderately correlated (r^2^=0.39).

Other identified loci share associations with several other traits such as social and emotional characteristics (e.g. *MMS22L* - *KLHL32*), lung function (e.g. *POM12IL2* - *PRSS16*), food/drink intake (e.g. *DARS1* - *CENPL*), and cognitive traits (e.g. *MDK* - *CHRM4*). However, our finding of an association of the *APOE* variant with *exercising alone* is likely the result of selection/survival bias due to older individuals with the ε4 risk allele being relatively enriched for healthy behaviors that has offset their genetic risk, and enabled their survival and participation in the study ^8^. It is possible that our estimates for other identified loci are subject to this bias, although it is likely to be minimal for nearly all genetic loci.

Although it turns out that we were under-powered to detect statistically significant associations in our replication cohorts, we did observe a nominally significant association of the *CADM2* variant with liking of *exercising with others* in the NTR, further reinforcing this locus directly or indirectly in PA behavior, in addition to other traits, as mentioned above. In PRS analyses, we found that these PA-liking measures were genetically consistent across the UK Biobank and NTR samples, and that they could contribute substantially to the prediction and genetic understanding of (at least) self-reported PA behavior. It should be noted that the proportion of phenotypic variance explained by these PRS is still extremely small. This is at least partly attributable to the degree of measurement error one would expect from any self-report measure.

The strengths of this study include the relatively large sample size, the multiple measures of PA-liking, the ability to examine correlations with both self-reported and accelerometry-derived PA behaviors, and the availability of two additional cohorts for replication and testing of PRS. The inclusion of middle-to-older-age adults of European-descent individuals is a limitation of our study as these results may not generalize to other groups.

In conclusion, we have identified genetic variants associated with PA-liking, further refining our understanding of the genetics of PA behavior. Our results show that PA-liking can capture additional elements of PA not captured by either self-report or accelerometry. Future work is needed to understand the mechanisms linking the identified loci to PA behavior, and how these may vary over the life-course. Examining the genetic correlates of stated liking for different facets of PA and how they associate with self-reported and device-based measures of PA can provide insight into both the genetic and non-genetic determinants of PA behavior, potentially help plan more effective interventions to improve PA habits, and bridge gaps that may exist between liking and engaging in PA.

## Supporting information

Supplemental Methods and Results

PheWas Results

## Data Availability

Summary statistics from genome-wide association studies presented in our study will be posted to the GWAS Catalog upon publication.

https://www.ukbiobank.ac.uk/

## ACKNOWLEDGMENTS

This research was conducted using the UK Biobank Resource under Application Number 15678 and 19655. The authors would like to thank the funders, organizers and participants of the UK Biobank, the Netherlands Twin Register, and TwinsUK. J.F.W. acknowledges support from the MRC Human Genetics Unit quinquennial programme grant “QTL in Health and Disease” (MC_UU_00007/10). TwinsUK receives funding from the Wellcome Trust (212904/Z/18/Z), Medical Research Council (AIMHY; MR/M016560/1) and European Union (H2020 contract #733100). TwinsUK and M.M. are supported by the National Institute for Health Research (NIHR)-funded BioResource, Clinical Research Facility and Biomedical Research Centre based at Guy’s and St Thomas’ NHS Foundation Trust in partnership with King’s College London. C.M. is funded by the Chronic Disease Research Foundation and by the Medical Research Council (MRC)/British Heart Foundation Ancestry and Biological Informative Markers for Stratification of Hypertension (AIMHY; MR/M016560/1). G.E.A. acknowledges support from the National Institute on Aging, NIH (AG072980, AG067200, AG064587), State of Arizona and Arizona Department of Health Services, and McKnight Brain Research Foundation. Phenotyping in NTR was funded by BBRMI-CP2011-38: Enrichment of NTR with information on dietary intake and aspects of eating behavior (PI: Boomsma & Feskens); genotyping in NTR was funded by multiple grants from the Netherlands Organization for Scientific Research (NWO) and The Netherlands Organisation for Health Research and Development (ZonMW); the Biobanking and Biomolecular Resources Research Infrastructure (BBMRI –NL, 184.021.007 and 184.033.111), the European Community’s Framework Programs (FP5-LIFE QUALITY-CT-2002-2006, FP7-HEALTH-F4-2007-2013, grant 01254: GenomEUtwin, grant 01413: ENGAGE); the European Research Council (ERC Starting 284167, ERC Consolidator 771057, ERC Advanced 230374), Rutgers University Cell and DNA Repository (NIMH U24 MH068457-06), the National Institutes of Health (NIH, R01D0042157-01A1, MH081802, DA018673, R01 DK092127-04, Grand Opportunity grant 1RC2 MH089951); and the Avera Institute for Human Genetics, Sioux Falls, South Dakota (USA).

## AUTHOR CONTRIBUTIONS

Conceived and designed study: YCK & NP. Performed data analyses: NP, YCK, AA, SMW, MN, VB, MM, CM, MDVZ, JJH & EJCdG. Drafting of manuscript: YCK & NP. Critically revised the manuscript for important intellectual content: YCK, NP, AA, SMW, MN, VB, MM, CM, DAR, GEA, MDVZ, JJH, JFW, EJCdG & DIB.

## DATA AVAILABILITY

Access to UK Biobank data is available to registered researchers upon application. GWAS summary statistics data for the UK Biobank PA-liking traits will be made available through the GWAS catalog portal at the time of publication.

