## Supplemental Methods and Results for "Genome-wide association study of liking of physical activity in the UK Biobank"

**Supplementary Methods**

**GWAS in Netherlands Twin Register (NTR)**

Genotyping was done on multiple platforms over time, namely Perlegen-Affymetrix, Affymetrix 6.0, Affymetrix Axiom, Illumina Human Quad Bead 660, Illumina Omni 1M and Illumina GSA. On each platform, genotyping was performed following manufacturers protocols, using the then appropriate calling software. For each genotype platform, samples were removed if DNA-based sex did not match the expected phenotype, if the PLINK heterozygosity F-statistic was < -0.10 or > 0.10, or if the genotyping call rate was < 0.90. SNPs were removed if the MAF < 10^-6^, if the Hardy-Weinberg equilibrium p-value was < 10^-6^, and/or if the call rate was < 0.95 ^1^. Subsequently, for each platform, the genotype data was aligned with the 1000 Genomes reference panel using the HRC and 1000 Genomes checking tool, which tests and filters for SNPs with allele frequency differences larger than 0.20 as compared to the CEU population as well as palindromic SNPs ^2,3^. DNA strand was flipped to the positive strand of the 1000 Genomes reference panel. The data of the six platforms was then merged into a single dataset, keeping all quality-controlled SNPs of each platform. For each individual, one platform was chosen. Based on the ~10.8k SNPs that all platforms have in common, DNA Identity By Descent state was estimated for all individual pairs using the PLINK v1.9 and KING programs ^4^. These estimates were then compared to the expected familial relations, and samples were removed if these failed to fit. CEU population outliers, based on per platform 1000 Genomes PC projection with the SMARTPCA software, were removed from the data ^5^. Then, per platform, the data was phased using EAGLE and then imputed to 1000 Genomes using MINIMAC3 following the Michigan imputation server protocols ^6^. Post imputation, the resulting separate platform Variant Call Format (VCF) files were merged with BCFTOOLS into a single VCF file per chromosome for each reference, only for those SNPs present on all six platforms ^7^. 10 genetic PCs were re-calculated with SMARTPCA using LD-pruned 1000 Genomes–imputed SNPs that were also genotyped on at least one platform, had MAF > 0.05 and were not present in the long-range LD regions, as described earlier ^8^.

**Polygenic scores (PGS) of PA-liking in NTR**

For the polygenic scoring the imputed data were converted to best guess genotype data and were filtered to include only ACGT SNPs, SNPs with MAF > 0.01, HWE p > 10^-5^, imputation quality R^2^ >=0.80 and a genotype call rate > 0.98. SNPs with more than 2 alleles at a certain chromosomal and base-pair location were also excluded. All mendelian errors were set to missing. Before calculating the PGSs, linkage disequilibrium (LD) adjusted beta coefficients were calculated from the summary statistics using the LDPRED package v1 to correct for the effects of LD and to maximize predictive accuracy of the PGSs ^9^. These beta coefficients were calculated employing a LD pruning window of 250 KB, with different cut-offs of the proportions of causal SNPs namely 0.001, 0.003, 0.005 0.01, 0.03, 0.05, 0.10, 0.20, 0.30, 0.50 and INF. 2500 unrelated genotyped people from the NTR were used as reference sample to estimate the LD between markers. Finally, the PGS for liking of *bicycling*, *exercising alone*, *exercising with others*, *going to the gym*, *working up a sweat*, were calculated using adjusted summary statistics beta coefficients with the PLINK v1.9 software.

Each prior of each PRS was regressed on each PA-liking item in the NTR using generalized estimating equation (GEE) models to correct for family-structure of the NTR data. Partial R^2^ values were obtained by subtracting the adjusted R^2^ of a model where PA liking item was predicted by the covariates (age, sex, 10 genetic PCs, and genotyping platform) from the adjusted R^2^ of a GEE model in which only the covariates were included.

**PRS of PA in NTR**

Participants in the NTR answered the following questions regarding regular exercise activities: (1) which activity/activities they engaged in; (2) the weekly frequency of each activity; (3) the average duration of each exercise bout (in minutes). Each exercise activity was then recoded to a metabolic equivalent of task (MET) ^10^. By taking the product of the average duration, the weekly frequency and MET value for each exercise a MET-minutes per week (METmin/week) value for each exercise activity is generated. Next by summing across all exercise activities within each individual a total METmin/wk score was generated.

We further classified all activities as belonging to team-based or solitary exercise activities. Team-based exercise activities are defined as being activities where a group of exercisers work together to reach a common goal or shared objective (e.g. football or hockey). Solitary exercise activities are activities that are always performed by one individual (e.g. fitness or running). A complete list of exercise activities that were defined as belonging to either team-based or solitary exercise is provided in Supplementary table 1 in van der Zee, et al. ^11^. For both team-based and solitary exercise METmin/week scores were generated similar to the total exercise score, with the difference that only exercise activities under the definition of either team-based or solitary exercise are included, and any exercise activities that do not meet the definition are counted as 0 for the respective scores.

A similar approach as mentioned in the previous PRS analysis was used to assess predictability of exercise behavior using three different exercise PRS (SSOE, MVPA, and fraction of acceleration > 425mG from ^12^). Note that exercise behavior was not (yet) available from the same survey as the PA liking items, therefore the mean age(s) of this sample deviate from those listed above. To get as close to the sample from the PA-liking items, the last completed survey for each participant was selected (as the PA liking item was from a more recent survey).

We added both the PRS of PA-liking and the PRS of exercise to assess if this could improve the prediction of exercise behavior. SSOE was used as exercise PRS, as it demonstrated much larger R^2^ compared to the other two across all exercise phenotypes. For PA-liking we included the top three (*exercising with others*, *going to the gym*, and GIP1) separately. From each PRS we used the most predictive prior for each exercise phenotype as determined in previous analyses.

**Supplementary Table 1:** Descriptive statistics of the UK Biobank, NTR and TwinsUK samples and PA-liking traits.

|  | **UKB** | **NTR** | **TwinsUK*** |
| --- | --- | --- | --- |
| **Sample size (n)** | 153,002 | 7,740 | 1,503 |
| **Age (years)** | 66.79 (7.63) | 45.01 (17.25) | 60.75 (10.69) |
| **% Female** | 57.01 | 66.58 | 91.55 |
| ***Going to the gym*; mean (sd)** | 4.67 (2.67) | 5.32 (2.89) | 0.46 (0.27) |
| ***Exercising alone*; mean (sd)** | 5.92 (2.29) | 6.19 (2.61) | 0.54 (0.26) |
| ***Exercising with others*; mean (sd)** | 6.01 (2.37) | 6.67 (2.6) | 0.64 (0.25) |
| ***Bicycling*; mean (sd)** | 5.56 (2.53) | 7.65 (1.88) | 0.51 (0.29) |
| ***Working up a sweat*; mean (sd)** | 5.84 (2.18) | 6.67 (2.25) | 0.55 (0.26) |

* TwinsUK used a continuous linear scale that was rescaled to a 0-1 scale.

**Supplementary Table 2:** Associations of age, sex, BMI, and socioeconomic status with PA- liking traits in the UK Biobank. The beta coefficient units refer to an increment of one on the 9-point liking scale.

|  | ***Bicycling*** | | |  | ***Exercising alone*** | | |  | ***Exercising with others*** | | |  | ***Going to the gym*** | | |  | ***Working up a sweat*** | | |
| --- | --- | --- | --- | --- | --- | --- | --- | --- | --- | --- | --- | --- | --- | --- | --- | --- | --- | --- | --- |
|  | **β** | **SE** | **p** |  | **β** | **SE** | **p** |  | **β** | **SE** | **p** |  | **β** | **SE** | **p** |  | **β** | **SE** | **p** |
| **Age** | -0.06 | 0 | 0 |  | -0.03 | 0 | 0 |  | -0.02 | 0 | 5.40E-97 |  | -0.03 | 0 | 7.30E-248 |  | -0.05 | 0 | 0 |
| **Sex (male)** | 1.13 | 0.01 | 0 |  | 0.72 | 0.01 | 0 |  | -0.44 | 0.01 | 9.00E-271 |  | 0.32 | 0.01 | 7.30E-105 |  | 1.04 | 0.01 | 0 |
| **BMI** | -0.37 | 0.01 | 0 |  | -0.35 | 0.01 | 0 |  | -0.31 | 0.01 | 0 |  | -0.17 | 0.01 | 5.90E-117 |  | -0.26 | 0.01 | 0 |
| **TDI** | 0 | 0.01 | 9.40E-01 |  | 0.01 | 0.01 | 3.30E-02 |  | -0.16 | 0.01 | 1.70E-128 |  | -0.05 | 0.01 | 7.30E-10 |  | -0.06 | 0.01 | 1.20E-29 |
| **College Degree** | 0.27 | 0.01 | 1.50E-92 |  | 0.23 | 0.01 | 6.00E-82 |  | -0.04 | 0.01 | 5.70E-04 |  | 0.02 | 0.02 | 1.60E-01 |  | -0.03 | 0.01 | 9.00E-03 |
| **Income (18-31k)** | 0.08 | 0.02 | 3.70E-04 |  | 0.01 | 0.02 | 5.50E-01 |  | 0.15 | 0.02 | 1.80E-11 |  | 0.16 | 0.03 | 7.20E-10 |  | 0.16 | 0.02 | 5.40E-16 |
| **Income (31-59k)** | 0.17 | 0.02 | 9.50E-15 |  | 0.07 | 0.02 | 4.10E-04 |  | 0.24 | 0.02 | 1.60E-29 |  | 0.34 | 0.03 | 1.10E-38 |  | 0.27 | 0.02 | 1.60E-47 |
| **Income (60k-100k)** | 0.29 | 0.02 | 5.90E-35 |  | 0.15 | 0.02 | 3.00E-13 |  | 0.35 | 0.02 | 1.10E-55 |  | 0.51 | 0.03 | 4.60E-79 |  | 0.42 | 0.02 | 9.60E-105 |
| **Income (>100k)** | 0.4 | 0.03 | 1.50E-41 |  | 0.25 | 0.03 | 9.10E-21 |  | 0.46 | 0.03 | 3.50E-56 |  | 0.83 | 0.03 | 5.30E-132 |  | 0.6 | 0.03 | 5.20E-127 |

p-value of 0 indicates p<2.2E-308; BMI: body mass index, TDI: Townsend deprivation index

**Supplementary Table 3:** Genomic inflation and heritability estimates from LD-score regression analysis in UK Biobank.

|  | **Lambda GC** | **Intercept** | **h^2^** |
| --- | --- | --- | --- |
| *Working up a sweat* | 1.184 | 0.9947 (0.0068) | 0.0711 (0.004) |
| *Going to the gym* | 1.1333 | 0.9929 (0.0066) | 0.0535 (0.004) |
| *Exercising with Others* | 1.1459 | 0.9984 (0.007) | 0.0571 (0.0039) |
| *Exercising Alone* | 1.1747 | 0.9949 (0.0072) | 0.0608 (0.0043) |
| *Bicycling* | 1.1908 | 1.0032 (0.0068) | 0.0754 (0.0038) |
| GIP1 | 1.1973 | 0.9777 (0.007) | 0.0887 (0.0042) |

h^2^: heritability; GC: genomic control.

**Supplementary Table 4:** Cell type enrichment based on overall PA-liking (GIP1) GWAS results (top 25 results according to p-value are shown).

| **Name** | **Coefficient** | **Coefficient_std_error** | **Coefficient_P_value** |
| --- | --- | --- | --- |
| Brain_Nucleus_accumbens_(basal_ganglia) | 7.47E-09 | 1.71E-09 | 6.39E-06 |
| Brain_Hippocampus | 6.42E-09 | 1.63E-09 | 4.18E-05 |
| A08.186.211.Brain | 6.40E-09 | 1.64E-09 | 4.80E-05 |
| Brain_Caudate_(basal_ganglia) | 6.30E-09 | 1.65E-09 | 6.90E-05 |
| Brain_Frontal_Cortex_(BA9) | 6.26E-09 | 1.67E-09 | 8.85E-05 |
| Brain_Amygdala | 5.84E-09 | 1.60E-09 | 1.29E-04 |
| Brain_Anterior_cingulate_cortex_(BA24) | 5.96E-09 | 1.71E-09 | 2.49E-04 |
| A08.186.211.464.Limbic.System | 6.35E-09 | 1.84E-09 | 2.71E-04 |
| A08.186.211.464.405.Hippocampus | 5.73E-09 | 1.68E-09 | 3.12E-04 |
| A08.186.211.464.710.225.Entorhinal.Cortex | 6.20E-09 | 1.86E-09 | 4.33E-04 |
| Brain_Putamen_(basal_ganglia) | 5.56E-09 | 1.69E-09 | 4.93E-04 |
| Brain_Hypothalamus | 5.42E-09 | 1.69E-09 | 6.88E-04 |
| A08.186.211.730.885.287.500.670.Parietal.Lobe | 5.65E-09 | 1.79E-09 | 8.04E-04 |
| Brain_Cortex | 5.28E-09 | 1.72E-09 | 1.06E-03 |
| A08.186.211.730.885.287.500.Cerebral.Cortex | 5.47E-09 | 1.82E-09 | 1.34E-03 |
| Brain_Substantia_nigra | 4.97E-09 | 1.66E-09 | 1.36E-03 |
| A08.186.211.730.885.287.500.571.735.Visual.Cortex | 4.50E-09 | 1.68E-09 | 3.66E-03 |
| Brain_Cerebellum | 4.44E-09 | 1.78E-09 | 6.35E-03 |
| Brain_Cerebellar_Hemisphere | 4.05E-09 | 1.66E-09 | 7.25E-03 |
| A08.186.211.730.885.287.249.Basal.Ganglia | 4.12E-09 | 1.74E-09 | 9.02E-03 |
| A06.407.071.140.Adrenal.Cortex | 3.57E-09 | 1.67E-09 | 1.61E-02 |
| A08.186.211.653.Mesencephalon | 3.97E-09 | 1.86E-09 | 1.65E-02 |
| A08.186.211.730.885.287.500.270.Frontal.Lobe | 3.66E-09 | 1.78E-09 | 1.97E-02 |
| A15.145.229.637.555.567.569.200.CD4.Positive.T.Lymphocytes | 4.16E-09 | 2.14E-09 | 2.59E-02 |
| A08.186.211.132.Brain.Stem | 3.33E-09 | 1.76E-09 | 2.92E-02 |

**Supplementary Table 5:** Correlations of PA-liking PRS (based on UK Biobank) with PA-liking phenotypes in NTR at different PRS priors for the proportion of causal SNPs.

|  |  | **Going to the gym** | | **Exercising alone** | | **Exercising together** | | **Bicycling** | | **Working up a sweat** | |
| --- | --- | --- | --- | --- | --- | --- | --- | --- | --- | --- | --- |
| **PRS** | **Prior** | **R2** | **p** | **R2** | **p** | **R2** | **p** | **R2** | **p** | **R2** | **p** |
| **Bicycling** | **P0001** | 0.00011 | 0.20883 | 7E-05 | 0.243554 | 0.00034 | 0.085628 | -6.9E-05 | 0.534264 | -0.000108 | 0.7630861 |
|  | **P0003** | 0.00243 | 2.5E-05 | 0.0026 | 2.08E-05 | 0.00341 | 8.82E-07 | 0.003 | 2.34E-06 | 0.001516 | 0.0005263 |
|  | **P0005** | 0.00173 | 0.00031 | 0.004 | 6.69E-08 | 0.00278 | 7.11E-06 | 0.00508 | 1.14E-09 | 0.001807 | 0.0001427 |
|  | **P001** | 0.00184 | 0.0002 | 0.0043 | 2.14E-08 | 0.00282 | 5.44E-06 | 0.00561 | 1.33E-10 | 0.001919 | 9.202E-05 |
|  | **P003** | 0.00192 | 0.00015 | 0.0046 | 7.54E-09 | 0.00273 | 6.99E-06 | 0.00572 | 7.22E-11 | 0.001986 | 7.106E-05 |
|  | **P005** | 0.00187 | 0.00019 | 0.0047 | 6.47E-09 | 0.00264 | 9.63E-06 | 0.00571 | 7.21E-11 | 0.001936 | 8.66E-05 |
|  | **P010** | 0.00188 | 0.00018 | 0.0047 | 6.53E-09 | 0.00265 | 9.12E-06 | 0.00573 | 6.38E-11 | 0.001954 | 8.089E-05 |
|  | **P020** | 0.00188 | 0.00018 | 0.0047 | 5.82E-09 | 0.00265 | 9.2E-06 | 0.00574 | 6.23E-11 | 0.001955 | 8.05E-05 |
|  | **P030** | 0.00188 | 0.00018 | 0.0047 | 5.76E-09 | 0.00264 | 9.28E-06 | 0.00574 | 6.03E-11 | 0.001946 | 8.361E-05 |
|  | **P050** | 0.00188 | 0.00018 | 0.0047 | 5.42E-09 | 0.00263 | 9.54E-06 | 0.00575 | 5.84E-11 | 0.00195 | 8.222E-05 |
|  | **INF** | 0.00185 | 0.0002 | 0.0047 | 7.02E-09 | 0.00261 | 1.07E-05 | 0.00587 | 3.67E-11 | 0.001974 | 7.286E-05 |
| **Exercising alone** | **P0001** | 0.00081 | 0.01037 | 0.0023 | 4.46E-05 | 0.00057 | 0.023416 | 0.0013 | 0.001563 | 0.001003 | 0.0034235 |
|  | **P0003** | 0.0029 | 5.2E-06 | 0.0073 | 9.72E-13 | 0.00114 | 0.002924 | 0.00534 | 5.7E-10 | 0.002019 | 6.147E-05 |
|  | **P0005** | 0.00254 | 2E-05 | 0.0069 | 4.1E-12 | 0.00099 | 0.0052 | 0.00496 | 2.69E-09 | 0.001738 | 0.0001994 |
|  | **P001** | 0.00234 | 4.1E-05 | 0.0064 | 1.68E-11 | 0.00085 | 0.008714 | 0.00455 | 1.18E-08 | 0.001596 | 0.0003626 |
|  | **P003** | 0.00215 | 8E-05 | 0.006 | 7.32E-11 | 0.00073 | 0.013695 | 0.0042 | 4.47E-08 | 0.001408 | 0.0007783 |
|  | **P005** | 0.00211 | 9.3E-05 | 0.0059 | 9.87E-11 | 0.00071 | 0.014505 | 0.0041 | 6.39E-08 | 0.001363 | 0.0009395 |
|  | **P010** | 0.00206 | 0.00011 | 0.0058 | 1.38E-10 | 0.00069 | 0.0161 | 0.00402 | 8.41E-08 | 0.001344 | 0.0010111 |
|  | **P020** | 0.00205 | 0.00012 | 0.0058 | 1.5E-10 | 0.00068 | 0.016361 | 0.00401 | 8.91E-08 | 0.001336 | 0.0010439 |
|  | **P030** | 0.00204 | 0.00012 | 0.0058 | 1.54E-10 | 0.00068 | 0.016599 | 0.004 | 9.13E-08 | 0.00133 | 0.0010709 |
|  | **P050** | 0.00204 | 0.00012 | 0.0058 | 1.65E-10 | 0.00068 | 0.016913 | 0.00399 | 9.61E-08 | 0.001328 | 0.0010763 |
|  | **INF** | 0.00202 | 0.00013 | 0.0057 | 2.41E-10 | 0.00066 | 0.018357 | 0.00403 | 8.21E-08 | 0.001343 | 0.0010057 |
| **Exercising with others** | **P0001** | -3E-05 | 0.40458 | 3E-05 | 0.284045 | 0.00011 | 0.202795 | -5.7E-05 | 0.504431 | -0.000117 | 0.8946558 |
|  | **P0003** | 0.00269 | 1.2E-05 | 0.0032 | 1.46E-06 | 0.00781 | 4.82E-13 | 0.00208 | 0.000128 | 0.004886 | 1.292E-09 |
|  | **P0005** | 0.0028 | 8.4E-06 | 0.0037 | 2.22E-07 | 0.00797 | 2.57E-13 | 0.00224 | 7.28E-05 | 0.005238 | 3.881E-10 |
|  | **P001** | 0.00275 | 1E-05 | 0.0041 | 5.79E-08 | 0.00791 | 2.7E-13 | 0.0024 | 4.17E-05 | 0.00551 | 1.719E-10 |
|  | **P003** | 0.00267 | 1.4E-05 | 0.0043 | 2.4E-08 | 0.00786 | 2.56E-13 | 0.00246 | 3.27E-05 | 0.005659 | 1.098E-10 |
|  | **P005** | 0.00266 | 1.4E-05 | 0.0043 | 1.78E-08 | 0.00785 | 2.57E-13 | 0.00249 | 2.93E-05 | 0.005671 | 1.053E-10 |
|  | **P010** | 0.00264 | 1.5E-05 | 0.0043 | 1.7E-08 | 0.00781 | 2.8E-13 | 0.00249 | 2.95E-05 | 0.005682 | 1.037E-10 |
|  | **P020** | 0.00263 | 1.6E-05 | 0.0044 | 1.54E-08 | 0.00778 | 3.08E-13 | 0.00249 | 2.87E-05 | 0.005669 | 1.078E-10 |
|  | **P030** | 0.00263 | 1.6E-05 | 0.0044 | 1.5E-08 | 0.00778 | 3.04E-13 | 0.00249 | 2.87E-05 | 0.005681 | 1.034E-10 |
|  | **P050** | 0.00262 | 1.6E-05 | 0.0044 | 1.53E-08 | 0.00776 | 3.26E-13 | 0.00249 | 2.86E-05 | 0.005675 | 1.069E-10 |
|  | **INF** | 0.00268 | 1.3E-05 | 0.0044 | 1.3E-08 | 0.00778 | 2.96E-13 | 0.00259 | 2.03E-05 | 0.005711 | 9.796E-11 |
| **Going to the gym** | **P0001** | 0.00141 | 0.00102 | 0.0018 | 0.000176 | 0.00232 | 3.05E-05 | 0.00066 | 0.01994 | 0.001858 | 0.0001409 |
|  | **P0003** | 0.00199 | 0.00012 | 0.0032 | 7.45E-07 | 0.00302 | 2.62E-06 | 0.00112 | 0.003278 | 0.003088 | 9.856E-07 |
|  | **P0005** | 0.00202 | 0.0001 | 0.0033 | 3.79E-07 | 0.00312 | 1.85E-06 | 0.00125 | 0.002003 | 0.003314 | 3.808E-07 |
|  | **P001** | 0.00211 | 7.6E-05 | 0.0037 | 9.08E-08 | 0.00325 | 1.15E-06 | 0.00136 | 0.001297 | 0.003622 | 1.027E-07 |
|  | **P003** | 0.00209 | 8.1E-05 | 0.0038 | 6.37E-08 | 0.00329 | 9.38E-07 | 0.00141 | 0.001016 | 0.003708 | 7.053E-08 |
|  | **P005** | 0.00209 | 8.3E-05 | 0.0038 | 5.28E-08 | 0.00329 | 9.44E-07 | 0.00143 | 0.000927 | 0.003725 | 6.593E-08 |
|  | **P010** | 0.00209 | 8.2E-05 | 0.0039 | 4.82E-08 | 0.00329 | 9.34E-07 | 0.00144 | 0.000904 | 0.003753 | 5.832E-08 |
|  | **P020** | 0.00209 | 8.3E-05 | 0.0039 | 5E-08 | 0.00329 | 9.34E-07 | 0.00144 | 0.000894 | 0.003757 | 5.713E-08 |
|  | **P030** | 0.00209 | 8.3E-05 | 0.0039 | 4.72E-08 | 0.00329 | 9.15E-07 | 0.00144 | 0.000885 | 0.003767 | 5.465E-08 |
|  | **P050** | 0.00209 | 8.3E-05 | 0.0039 | 4.68E-08 | 0.00329 | 9.16E-07 | 0.00144 | 0.00088 | 0.003771 | 5.393E-08 |
|  | **INF** | 0.00214 | 6.8E-05 | 0.0039 | 3.87E-08 | 0.00337 | 6.85E-07 | 0.00147 | 0.000766 | 0.00376 | 5.452E-08 |
| **Working up a sweat** | **P0001** | 5E-05 | 0.28354 | -1E-05 | 0.374545 | -5E-05 | 0.480551 | -7.1E-05 | 0.547163 | -0.000115 | 0.8613689 |
|  | **P0003** | 0.00266 | 1.1E-05 | 0.0049 | 1.75E-09 | 0.00295 | 2.13E-06 | 0.00371 | 4.09E-07 | 0.004313 | 1.48E-08 |
|  | **P0005** | 0.003 | 2.7E-06 | 0.0051 | 7.98E-10 | 0.00298 | 2E-06 | 0.00343 | 1.08E-06 | 0.004431 | 1.183E-08 |
|  | **P001** | 0.00304 | 2.1E-06 | 0.005 | 1.13E-09 | 0.00281 | 3.96E-06 | 0.003 | 4.85E-06 | 0.004338 | 2E-08 |
|  | **P003** | 0.00313 | 1.4E-06 | 0.0049 | 1.66E-09 | 0.00276 | 5.39E-06 | 0.00276 | 1.08E-05 | 0.00425 | 3.285E-08 |
|  | **P005** | 0.00316 | 1.3E-06 | 0.0048 | 1.75E-09 | 0.00272 | 6.11E-06 | 0.00274 | 1.2E-05 | 0.004229 | 3.676E-08 |
|  | **P010** | 0.00313 | 1.4E-06 | 0.0048 | 2.28E-09 | 0.00269 | 7.01E-06 | 0.00267 | 1.51E-05 | 0.004188 | 4.431E-08 |
|  | **P020** | 0.00313 | 1.4E-06 | 0.0048 | 2.36E-09 | 0.00267 | 7.65E-06 | 0.00265 | 1.61E-05 | 0.004169 | 4.777E-08 |
|  | **P030** | 0.00314 | 1.3E-06 | 0.0048 | 2.4E-09 | 0.00268 | 7.35E-06 | 0.00265 | 1.64E-05 | 0.004163 | 4.904E-08 |
|  | **P050** | 0.00315 | 1.3E-06 | 0.0048 | 2.45E-09 | 0.00267 | 7.73E-06 | 0.00263 | 1.71E-05 | 0.004162 | 4.945E-08 |
|  | **INF** | 0.00321 | 1E-06 | 0.0048 | 2.4E-09 | 0.00273 | 6.1E-06 | 0.00271 | 1.31E-05 | 0.004196 | 4.242E-08 |
| **GIP1** | **P0001** | 7.7E-05 | 0.23484 | 4E-05 | 0.275032 | 0.0003 | 0.086494 | 6.5E-05 | 0.246366 | 2.94E-05 | 0.2981008 |
|  | **P0003** | 0.00011 | 0.20176 | 0.0016 | 0.000615 | 0.00083 | 0.008559 | 0.00104 | 0.004391 | 0.000951 | 0.0039202 |
|  | **P0005** | 0.00371 | 2.1E-07 | 0.0066 | 4.6E-12 | 0.00508 | 2.34E-09 | 0.00462 | 1.84E-08 | 0.004891 | 1.08E-09 |
|  | **P001** | 0.00402 | 6.4E-08 | 0.0074 | 1.85E-13 | 0.0054 | 5.88E-10 | 0.00505 | 3.46E-09 | 0.005425 | 1.65E-10 |
|  | **P003** | 0.00413 | 3.8E-08 | 0.0081 | 1.58E-14 | 0.00549 | 3.46E-10 | 0.00523 | 1.66E-09 | 0.005716 | 6.023E-11 |
|  | **P005** | 0.00417 | 3.3E-08 | 0.0081 | 1.25E-14 | 0.00552 | 2.9E-10 | 0.00527 | 1.43E-09 | 0.005748 | 5.436E-11 |
|  | **P010** | 0.00413 | 3.8E-08 | 0.0082 | 1.04E-14 | 0.0055 | 3.02E-10 | 0.00527 | 1.4E-09 | 0.005767 | 5.087E-11 |
|  | **P020** | 0.00415 | 3.5E-08 | 0.0082 | 8.41E-15 | 0.00549 | 3.08E-10 | 0.00525 | 1.47E-09 | 0.005784 | 4.838E-11 |
|  | **P030** | 0.00416 | 3.4E-08 | 0.0082 | 7.6E-15 | 0.00551 | 2.88E-10 | 0.00528 | 1.38E-09 | 0.005809 | 4.417E-11 |
|  | **P050** | 0.00416 | 3.3E-08 | 0.0082 | 7.61E-15 | 0.0055 | 2.95E-10 | 0.00527 | 1.41E-09 | 0.005805 | 4.533E-11 |
|  | **INF** | 0.00421 | 2.8E-08 | 0.0083 | 7.03E-15 | 0.00556 | 2.3E-10 | 0.00546 | 7.13E-10 | 0.005837 | 3.892E-11 |

**Supplementary Figure 1:** Prediction of self-reported PA in NTR by PA and PA-liking PRS derived from UK Biobank.

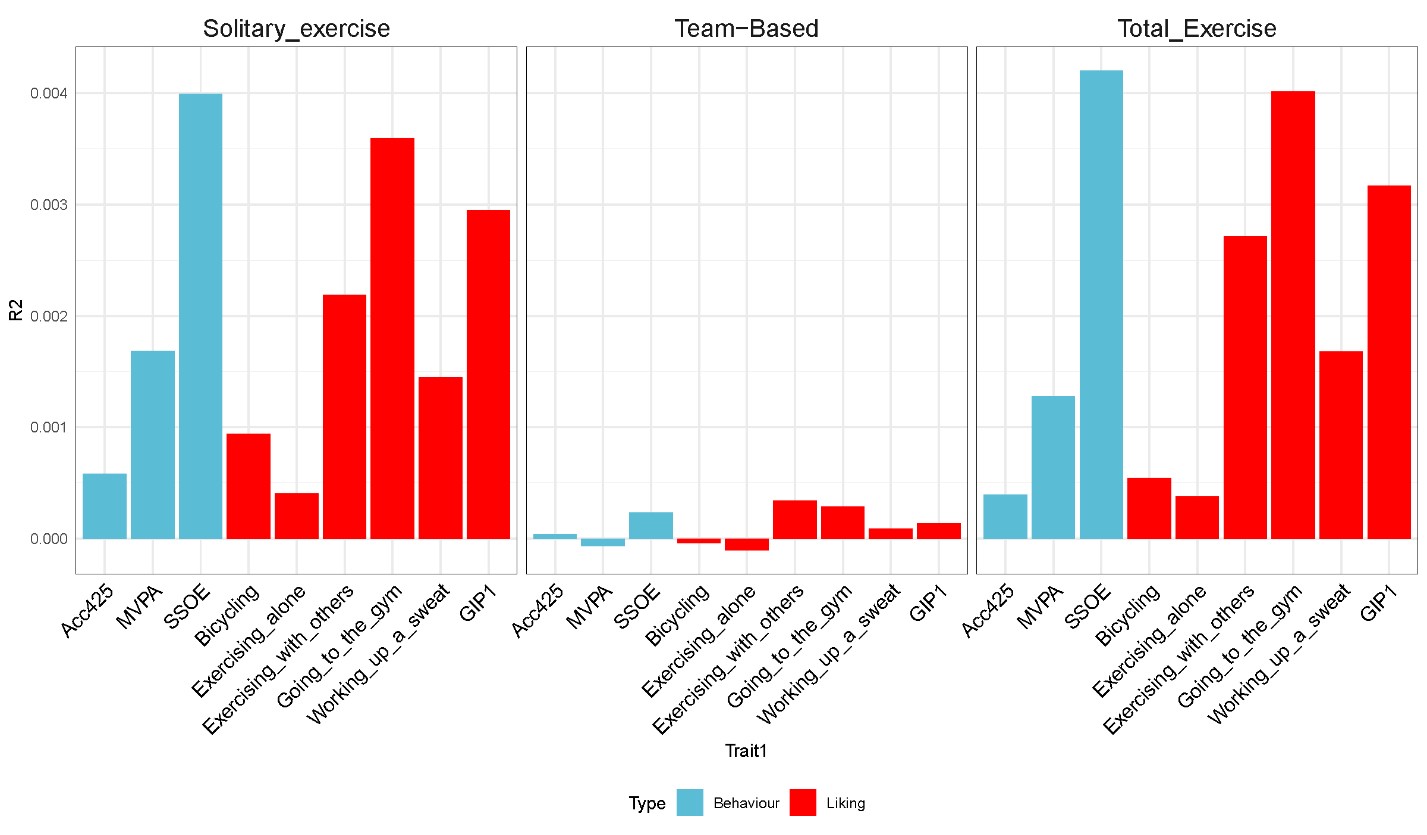

**Supplementary Figure 2:** Added prediction of self-reported PA in NTR by PA-liking PRSs over and beyond SSOE PRS.

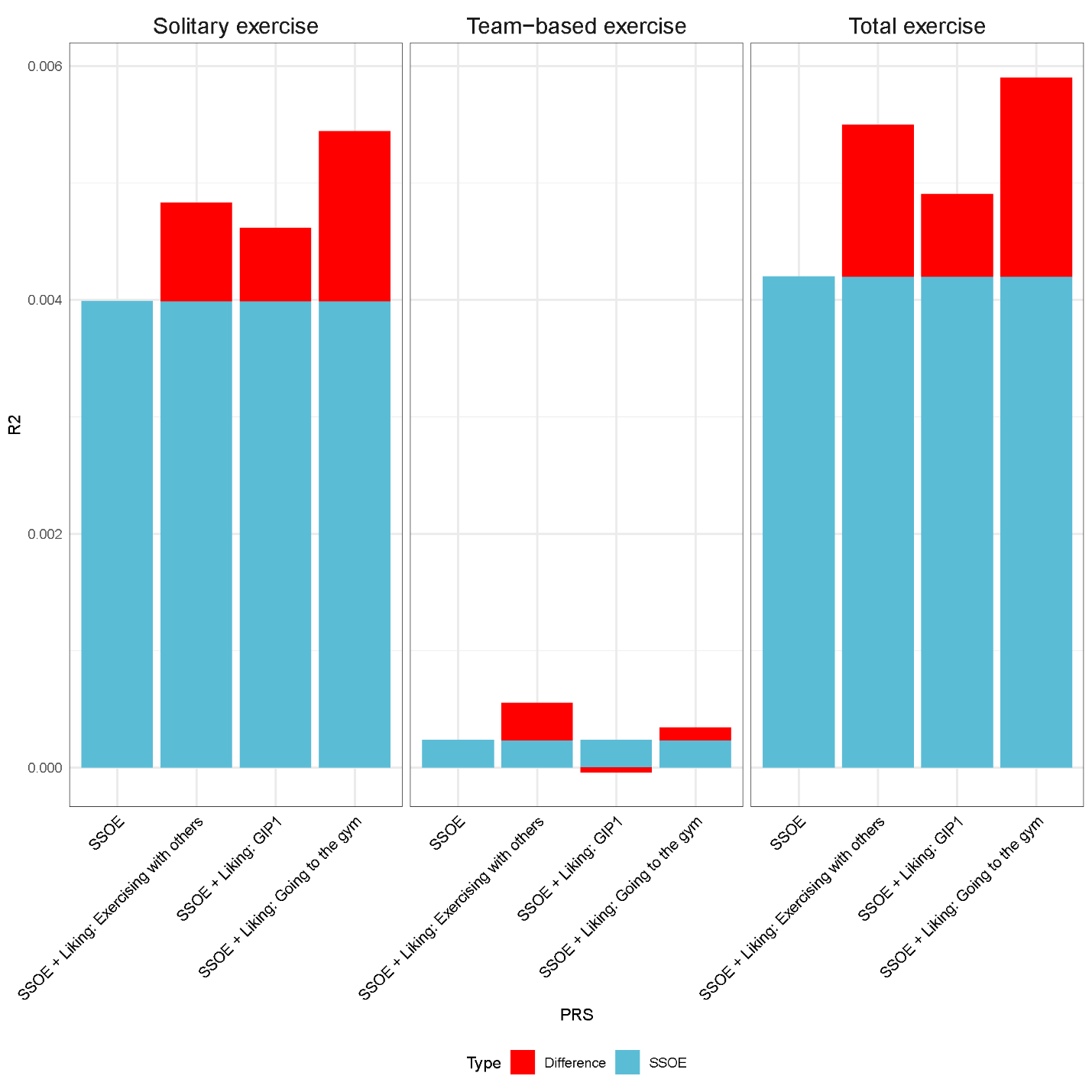
